# The effect of an incentive billing code on heart failure management in primary care: a population-based study

**DOI:** 10.1101/2024.08.16.24312144

**Authors:** Shijie Zhou, Douglas S. Lee, Francis Nguyen, Harsukh Benipal, Richard Perez, Peter C. Austin, Husam Abdel-Qadir, Jacob A. Udell, Catherine Demers

## Abstract

**Background:** To support family physicians (FPs) in managing patients with heart failure (HF), the Ministry of Health in Ontario, Canada, implemented the Q050 billing code in 2008, a pay-for-performance (P4P) incentive for guideline-based HF care. We studied whether the incentive was associated with any change in process-of-care measure, particularly the prescriptions of HF medications.

**Methods:** We identified all patients with HF in Ontario of age≥66, who were managed by FPs claiming the Q050 incentive between 2008 and 2021. We counted the number of patients who were prescribed renin-angiotensin system inhibitors (RASi), beta-blockers (BB), mineralocorticoid receptor antagonists (MRA), and diuretics three months before and after the Q050 billing code was claimed for these patients. Where applicable, we classified the agents within each class by whether they are guideline-directed as recommended by the Canadian Cardiovascular Society (CCS).

**Results:** We included 39,425 HF patients in the study. The median age was 80 (IQR 73-85) years; 49% were female. Half were assessed by an internist or cardiologist during the six months before their HF diagnosis. Compared to pre-Q050, there was an increase in RASi prescriptions from 42.5% to 45.8%, BB from 51.9% to 54.4%, MRA from 9.2% to 11.7%, and diuretics from 63.2% to 65.7% after the incentive (p<0.05). There was a decrease in those not on any HF medications from 27.5% to 24.9% (p<0.001). Those with newly diagnosed HF and prompt follow-up with FPs experienced the largest but clinically modest increase in HF medications.

**Conclusions:** To our knowledge, this is the first evaluation of process-of-care measures related to a pay-for-performance program in primary care HF management. The Q050 incentive led to a minimal increase in the prescription of HF medications; there is underutilization of disease-modifying agents. Further research is needed to understand why pay-for-performance programs had no effect on physician prescribing behaviours.

## Introduction

Heart failure (HF) presents an important healthcare burden; 50,000 new cases are diagnosed each year in Canada alone ^1^. Comprehensive HF care is shown to reduce HF hospitalizations, mortality, and healthcare costs ^2,3^.

Family physicians (FPs) play a crucial role in providing primary care and long-term HF management ^4^. To incentivize FPs to provide comprehensive guideline-based HF care, the Ontario Ministry of Health implemented a pay-for-performance (P4P) program in 2008 in the form of an incentive billing code, Q050. This is a lump-sum payment that can be claimed by FPs annually for HF management, including routine physical exams, optimization of HF medications, and completion of diagnostic tests.

Among patients managed by FPs who received the Q050 payments, a prior study demonstrated that there was no reduction in HF hospitalizations, emergency visits, or deaths ^5^. In the current analysis, we aimed to further investigate underlying processes of care measure, specifically in the realm of pharmacotherapy. Thus, we examined whether the billing code was associated with changes in the prescriptions of HF medications, including disease-modifying agents (renin-angiotensin system inhibitors, beta-blockers, mineralocorticoid receptor antagonists) and decongestive therapies (diuretics). We hypothesized that the incentive program was associated with no or a small increase in the prescription rates of HF medications.

## Methods

### The Q050A Billing Code

Introduced in April 2008, the Heart Failure Management Incentive billing code Q050A is a $125 CAD annual payment that can be claimed by family physicians for managing patients with HF in the outpatient setting. To be eligible to claim the code, family physicians are expected to coordinate and document essential elements of HF care in accordance with the HF guideline of the Canadian Cardiovascular Society. The required HF care involves: 1) comprehensive physical examination 2) laboratory monitoring of serum sodium, potassium, creatinine, and estimated glomerular filtration rate (eGFR) 3) patient education for modifiable risk factor reduction and self-management 4) pharmacologic management for appropriate use of first-line, symptom relief, and preventative medications. This process requires a minimum of two patient visits to complete.

### Study design and patient selection

This was a retrospective before-after cohort study conducted in Ontario, Canada; we compared the outcome before and after the billing of the Q050 incentive code. From the Institute of Clinical Evaluative Sciences (ICES) Congestive Heart Failure database, we identified all patients aged 66 years or older who were diagnosed with heart failure (HF) between 2008 and 2021. We included patients whose family physicians billed the Q050A code within one year of HF diagnosis to capture the initial medication changes in response to a HF diagnosis.

Exclusion criteria included patients: 1) who were non-Ontario residents at the time of HF diagnosis 2) whose family physicians first billed the Q050A code more than one year after their HF diagnosis 3) not eligible for the Ontario Health Insurance Plan (OHIP) at the time of HF diagnosis 4) who received palliative care within 6 months prior to HF diagnosis 5) with a cardiac transplant prior to HF diagnosis 6) who died before the first billing date 7) who were in long-term care facilities during the 6 months prior to HF diagnosis.

### Data sources

The datasets from this study is held securely in coded form at ICES. ICES is an independent, non-profit research institute funded by an annual grant from the Ontario Ministry of Health (MOH) and Ministry of Long-Term Care (MLTC). As a prescribed entity under Ontario’s privacy legislation, ICES is authorized to collect and analyse health care and demographic data, without consent, for health system evaluation and improvement.

We identified patients with heart failure (HF) from the ICES Congestive Heart Failure database, which identifies patients within the population with a sensitivity of 84.8% and specificity of 97.0% ^6^. A patient is considered to have HF if there is either one hospital record related to HF or one ambulatory record for HF followed by a second HF record from any source within one year. The date of a patient’s entry into the CHF database was used as an indicator of time of HF diagnosis. We obtained medication information from the Ontario Drug Benefit database (ODB), which records dispensation of all ODB-covered medications including those of interest to our study. Since all OHIP-covered Ontario residents of age ≥ 65 years are covered under the ODB, the database sufficiently captured medication prescriptions among study patients. Baseline demographics and comorbidities were obtained from the Canadian Institute for Health Information Discharge Abstract Database or from derived ICES datasets (the Ontario Diabetes Database, the Hypertension database, and the Chronic Obstructive Pulmonary Disease Database) or from OHIP billing records with relevant ICD and diagnostic codes as previously published ^7^. There was minimal missing data in the administrative databases. These datasets were linked using unique encoded identifiers and analyzed at ICES (formerly the Institute for Clinical Evaluative Sciences).

### Data sharing statement

The datasets from this study are held securely in coded form at the ICES. Although legal data-sharing agreements prohibit ICES from making the data set publicly available, access may be granted to those who meet prespecified criteria for confidential access, available at https://www.ices.on.ca/DAS. The full dataset creation plan and underlying analytic code are available from the authors upon request, understanding that the computer programs may rely on coding templates or macros that are unique to ICES and are, therefore, inaccessible or may require modification.

### Outcomes

The primary outcome was the prescription of each class of HF medications: renin-angiotensin system inhibitor (RASi), beta-blockers (BB), mineralocorticoid receptor antagonist (MRA), and diuretics.

RASi included angiotensin converting enzyme inhibitors (ACEi), angiotensin receptor blocker (ARB), and angiotensin receptor/neprilysin inhibitors (ARNI). We further differentiated the RASi and BB by whether the drug was evidence-based and directed by the CCS HF guideline^8^; we did not differentiate MRAs, as all MRAs on ODB formulary were guideline-directed. Sodium-glucose cotransporters-2 (SGLT2) inhibitors were not studied, as they were not considered contemporary therapies during majority of the study period.

### Subgroup analysis

We stratified our study population into four subgroups. Since those with HF diagnosed in hospital – often during acute heart failure – were a higher risk population compared to outpatient incident cases ^9^, we differentiated the population between inpatient and outpatient diagnoses. We further stratified the groups by the timing of Q050 billing at less or more than 3 months after HF diagnosis.

### Statistical analysis

Baseline characteristics and outcomes were summarized and compared among the four subgroups. Categorical variables were displayed as counts (percentage); continuous variables were summarized as median (interquartile range) and/or mean (standard deviation). We compared the proportion of patients with prescriptions of interest during the three months before and after the Q050A code was billed. Statistical significance in outcomes was calculated using the McNemar’s test given the matched nature of the comparison.

All analyses were performed using SAS version 9.4 (SAS Institute Inc., Cary, NC). The statistical significance of outcome comparisons between groups was defined as a two-tailed p-value <0.05. Cells with <6 individuals are suppressed to reduce the risk of re-identification as per ICES contractual obligations with data providers.

## Results

We included 39,245 patients in our study (figure 1). The median age at HF diagnosis was 80 (IQR 73-85) years; 48.9% percent were female. The most common cardiac comorbidities were hypertension (88.2%), diabetes (41.6%), and atrial fibrillation (19.1%). Only 12.4% were rural residents. There was high utilization of primary care resources: the mean number of primary care visits per year during the 5 years preceding HF diagnosis was 6.5. Half were also assessed by either an internist or cardiologist during the 6 months before their HF diagnosis (table 1).

**Figure 1:**
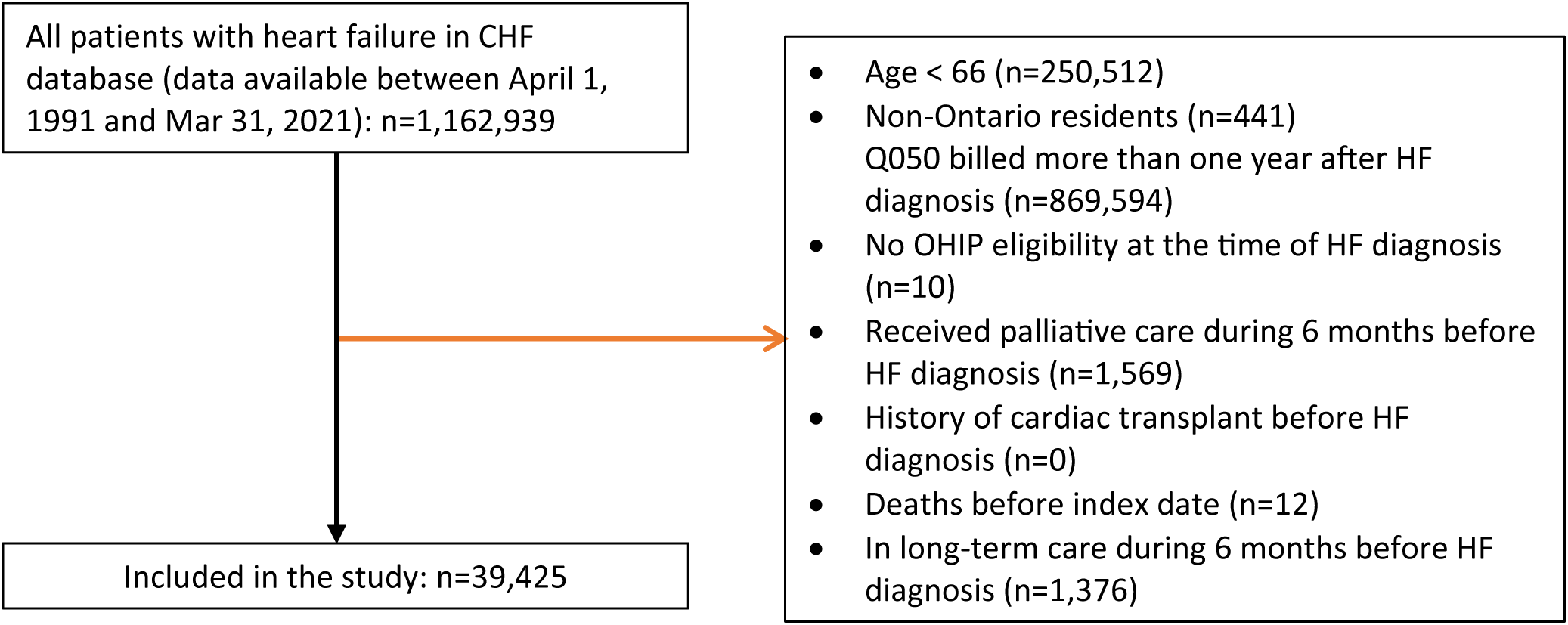

**Table 1:**
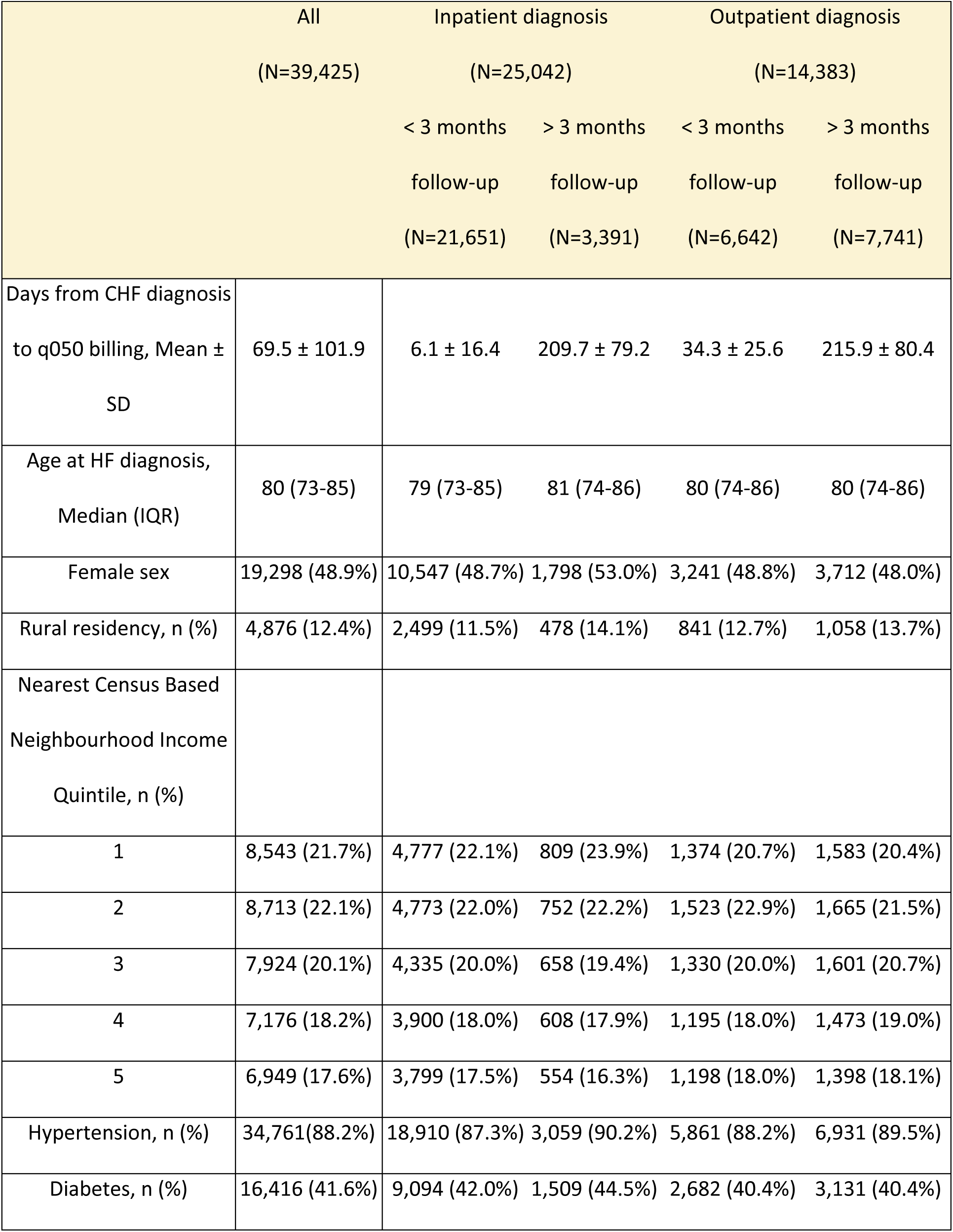
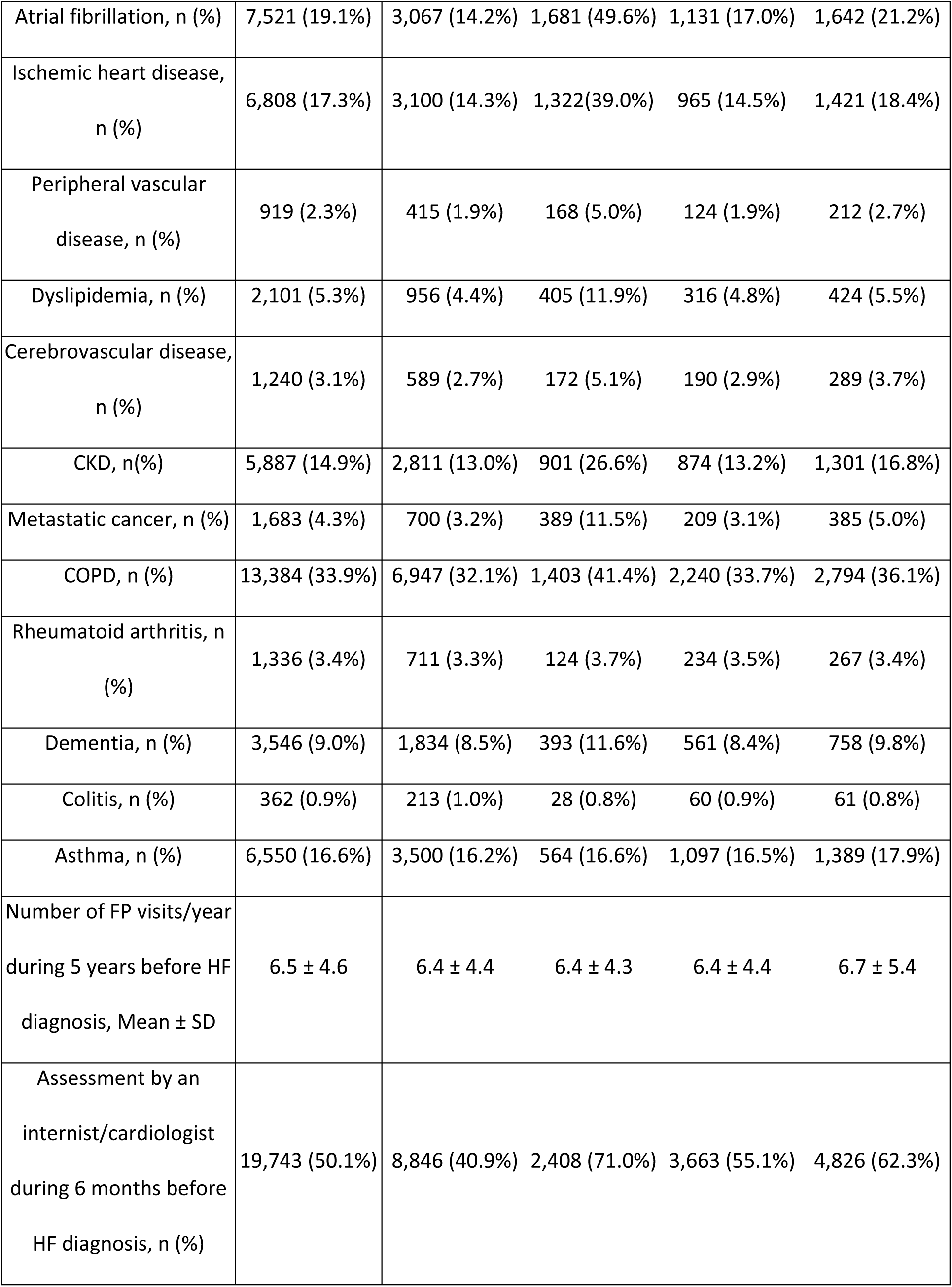
Baseline characteristics of study population.

|  | All | Inpatient diagnosis |  | Outpatient diagnosis |  |
| --- | --- | --- | --- | --- | --- |
|  | (N=39,425) | (N=25,042) |  | (N=14,383) |  |
|  |  | < 3 months | > 3 months | < 3 months | > 3 months |
|  |  | follow-up | follow-up | follow-up | follow-up |
|  |  | (N=21,651) | (N=3,391) | (N=6,642) | (N=7,741) |
| Days from CHF diagnosis to q050 billing, Mean ± SD | 69.5 ± 101.9 | 6.1 ± 16.4 | 209.7 ± 79.2 | 34.3 ± 25.6 | 215.9 ± 80.4 |
| Age at HF diagnosis, Median (IQR) | 80 (73-85) | 79 (73-85) | 81 (74-86) | 80 (74-86) | 80 (74-86) |
| Female sex | 19,298 (48.9%) | 10,547 (48.7%) | 1,798 (53.0%) | 3,241 (48.8%) | 3,712 (48.0%) |
| Rural residency, n (%) | 4,876 (12.4%) | 2,499 (11.5%) | 478 (14.1%) | 841 (12.7%) | 1,058 (13.7%) |
| Nearest Census Based Neighbourhood Income Quintile, n (%) |  |  |  |  |  |
| 1 | 8,543 (21.7%) | 4,777 (22.1%) | 809 (23.9%) | 1,374 (20.7%) | 1,583 (20.4%) |
| 2 | 8,713 (22.1%) | 4,773 (22.0%) | 752 (22.2%) | 1,523 (22.9%) | 1,665 (21.5%) |
| 3 | 7,924 (20.1%) | 4,335 (20.0%) | 658 (19.4%) | 1,330 (20.0%) | 1,601 (20.7%) |
| 4 | 7,176 (18.2%) | 3,900 (18.0%) | 608 (17.9%) | 1,195 (18.0%) | 1,473 (19.0%) |
| 5 | 6,949 (17.6%) | 3,799 (17.5%) | 554 (16.3%) | 1,198 (18.0%) | 1,398 (18.1%) |
| Hypertension, n (%) | 34,761(88.2%) | 18,910 (87.3%) | 3,059 (90.2%) | 5,861 (88.2%) | 6,931 (89.5%) |
| Diabetes, n (%) | 16,416 (41.6%) | 9,094 (42.0%) | 1,509 (44.5%) | 2,682 (40.4%) | 3,131 (40.4%) |
| Atrial fibrillation, n (%) | 7,521 (19.1%) | 3,067 (14.2%) | 1,681 (49.6%) | 1,131 (17.0%) | 1,642 (21.2%) |
| Ischemic heart disease, n (%) | 6,808 (17.3%) | 3,100 (14.3%) | 1,322 (39.0%) | 965 (14.5%) | 1,421 (18.4%) |
| Peripheral vascular disease, n (%) | 919 (2.3%) | 415 (1.9%) | 168 (5.0%) | 124 (1.9%) | 212 (2.7%) |
| Dyslipidemia, n (%) | 2,101 (5.3%) | 956 (4.4%) | 405 (11.9%) | 316 (4.8%) | 424 (5.5%) |
| Cerebrovascular disease, n (%) | 1,240 (3.1%) | 589 (2.7%) | 172 (5.1%) | 190 (2.9%) | 289 (3.7%) |
| CKD, n(%) | 5,887 (14.9%) | 2,811 (13.0%) | 901 (26.6%) | 874 (13.2%) | 1,301 (16.8%) |
| Metastatic cancer, n (%) | 1,683 (4.3%) | 700 (3.2%) | 389 (11.5%) | 209 (3.1%) | 385 (5.0%) |
| COPD, n (%) | 13,384 (33.9%) | 6,947 (32.1%) | 1,403 (41.4%) | 2,240 (33.7%) | 2,794 (36.1%) |
| Rheumatoid arthritis, n (%) | 1,336 (3.4%) | 711 (3.3%) | 124 (3.7%) | 234 (3.5%) | 267 (3.4%) |
| Dementia, n (%) | 3,546 (9.0%) | 1,834 (8.5%) | 393 (11.6%) | 561 (8.4%) | 758 (9.8%) |
| Colitis, n (%) | 362 (0.9%) | 213 (1.0%) | 28 (0.8%) | 60 (0.9%) | 61 (0.8%) |
| Asthma, n (%) | 6,550 (16.6%) | 3,500 (16.2%) | 564 (16.6%) | 1,097 (16.5%) | 1,389 (17.9%) |
| Number of FP visits/year during 5 years before HF diagnosis, Mean ± SD | 6.5 ± 4.6 | 6.4 ± 4.4 | 6.4 ± 4.3 | 6.4 ± 4.4 | 6.7 ± 5.4 |
| Assessment by an internist/cardiologist during 6 months before HF diagnosis, n (%) | 19,743 (50.1%) | 8,846 (40.9%) | 2,408 (71.0%) | 3,663 (55.1%) | 4,826 (62.3%) |

Of the four subgroups, those diagnosed as inpatient with a >3-month follow-up was the smallest with only 3,391 patients compared to 21,651, 6,642, and 7,741 in the other subgroups (Table 3). It also had the highest rates of comorbidities compared to other subgroups as well as the overall cohort: hypertension was present in 90.2%, diabetes in 44.5%, and atrial fibrillation in 49.6%; 71% were managed by an internist or cardiologist prior to their HF diagnosis, compared to 50% in the overall study (table 1).

Marginally more patients were prescribed each class of HF medications during the three months after the Q050 billing compared to the three months before. There was an increase from 45.2% to 45.8% in RASi (p=0.001), 51.9% to 54.4% in BB (p<0.0001), 9.2% to 11.7% in MRA (p<0.0001), and 63.2% to 65.7% in diuretic prescriptions (p<0.0001). Beta-blocker and MRA prescriptions increased the most, albeit by 2.5 percentage point only (table 2). The mean number of HF medications prescribed increased from 1.7 to 1.8 after Q050 was billed. Meanwhile, there was a small decrease from 27.5% to 24.9% in patients not on any disease-modifying agents such as RASi, BB or MRA (p<0.001). Of all RASi prescriptions, the proportion of guideline-directed agents increased from 66.8% to 67.9% (p<0.001); a similar increase from 84.3% to 85.6% was seen among the beta-blocker prescriptions (table 2).

**Table 2:** Number and proportions of patients in the entire cohort on each class of HF medications.

| All (N=39,425) |  |  | p-value |
| --- | --- | --- | --- |
|  | Pre-Q050 | Post-Q050 |  |
| Any RASi, n (%) | 17,827 (45.2%) | 18,066 (45.8%) | <0.05 |
| % guideline-directed | 66.8% | 67.9% | <0.001 |
| Any beta-blocker (n,%) | 20,477 (51.9%) | 21,431 (54.4%) | <0.001 |
| % guideline-directed | 84.3% | 85.6% | <0.001 |
| Any MRA | 3,645 (9.2%) | 4,608 (11.7%) | <0.001 |
| Any diuretics | 24,897 (63.2%) | 25,901 (65.7%) | <0.001 |
| % loop diuretics | 85.3% | 87.2% | <0.001 |
| Mean number of HF medications | 1.7 | 1.8 |  |
| Not on any HF medications | 6,061 (15.4%) | 5,049 (12.8%) | <0.001 |
| Not on any RASi, BB, or MRA | 10,834 (27.5%) | 9,804 (24.9%) | <0.001 |

Sixty-four percent were diagnosed with HF as inpatients, 36% as outpatients. Most inpatient incident cases were assessed by their FPs within 3 months of HF diagnosis (table 3). This was the largest subgroup which comprised 55% of the total study population; it also had the lowest rate of co-management by an internist or cardiologist during the six months preceding their HF diagnosis (40.9%) (table 1). Medication changes around the time of Q050 billing in this subgroup were more likely related to a new HF diagnosis and initiated by family physicians, therefore representing the most sensitive pre-post comparisons. Indeed, this was reflected in the much lower rates of BB (46.2% vs 69.3%, 56.6%, 56.3%, p<0.001), MRA (5.7% vs 21.3%, 10.3%, 13.0%, p<0.001), and diuretic (52.4% vs 80.1, 82.7%, 69.2%, p<0.001) prescriptions in this subgroup before Q050 billing. After Q050A, all classes of medications increased and were statistically significant. Diuretic saw the largest absolute increase of 6.8%, followed by BB (3.2%), MRA (2.6%), and RASi (1.5%). The proportion of patients not on any HF medications or on any disease-modifying agents (RASi, BB, MRA) saw an absolute decrease from 21.7% to 16.3% and from 31.9% to 28% respectively, albeit they were still the highest among all subgroups (table 3).

**Table 3:** Number and proportions of patients in each subgroup on each class of HF medications.

|  | Inpatient diagnosis<br>(N=25,042) |  |  |  | Outpatient diagnosis<br>(N=14,383) |  |  |  |
| --- | --- | --- | --- | --- | --- | --- | --- | --- |
|  | < 3 months follow-up<br>(N=21,651) |  | > 3 months follow-up<br>(N=3,391) |  | < 3 months follow-up<br>(N=6,642) |  | > 3 months follow-up<br>(N=7,741) |  |
|  | Pre-Q050 | Post-Q050 | Pre-Q050 | Post-Q050 | Pre-Q050 | Post-Q050 | Pre-Q050 | Post-Q050 |
| Any RASi, n (%) | 9,576<br>(44.2%)<br>** | 9,904<br>(45.7%)<br>** | 1,392<br>(41.0%) | 1,392<br>(41.0%) | 3,268<br>(49.2%) | 3,222<br>(48.5%) | 3,591<br>(46.4%) | 3,548<br>(45.8%) |
| % guideline-directed | 64.5%<br>** | 65.8%<br>** | 73.8% | 73.9% | 67.5% | 69.5% | 69.9% | 69.8% |
| Any beta-blocker, n (%) | 10,006<br>(46.2%)<br>** | 10,690<br>(49.4%)<br>** | 2,351<br>(69.3%) | 2,313<br>(68.2%) | 3,760<br>(56.6%)<br>** | 3,978<br>(59.9%)<br>** | 4,360<br>(56.3%) | 4,450<br>(57.5%) |
| % guideline-directed | 80.7%<br>** | 82.7%<br>** | 92.5% | 92.4% | 84.3%<br>** | 86.5%<br>** | 87.9%<br>* | 88.3%<br>* |
| Any MRA, n (%) | 1,233<br>(5.7%)<br>** | 1,806<br>(8.3%)<br>** | 722<br>(21.3%) | 717<br>(21.1%) | 684<br>(10.3%)<br>** | 962<br>(14.5%)<br>** | 1,006<br>(13.0%)<br>** | 1,123<br>(14.5%)<br>** |
| Any diuretics, n (%) | 11,336<br>(52.4%)<br>** | 12,828<br>(59.2%)<br>** | 2,715<br>(80.1%)<br>* | 2,641<br>(77.9%)<br>* | 5,493<br>(82.7%)<br>** | 5,071<br>(76.3%)<br>** | 5,353<br>(69.2%) | 5,361<br>(69.3%) |
| % loop diuretics | 75.4%<br>** | 81.2%<br>** | 97.2%<br>* | 97.2%<br>* | 93.9%<br>** | 93.4%<br>** | 91.5% | 91% |
| Average number<br>of HF<br>medications | 1.5 | 1.6 | 2.1 | 2.1 | 2.0 | 2.0 | 1.9 | 1.9 |
| Not on any HF<br>medications | 4,689<br>(21.7%)<br>** | 3,534<br>(16.3%)<br>** | 190<br>(5.6%) | 231(6.8%) | 374<br>(5.6%) | 496<br>(7.5%) | 808<br>(10.4%) | 788 (10.2%) |
| Not on any RASi,<br>BB, or MRA | 6,903<br>(31.9%)<br>** | 5,766<br>(28%)<br>** | 597<br>(17.6%) | 617<br>(18.2%) | 1514<br>(22.8%)<br>** | 1,343<br>(20.4%)<br>** | 1820<br>(23.5%) | 1,768<br>(22.9%) |

\*represents a p-value of <0.05 when pre- and post-Q050 rates were compared. \*\*represents a p-value of <0.001 when pre- and post-Q050 rates were compared.

Those diagnosed outpatient and assessed within 3 months of diagnosis also experienced an increase in beta-blocker and MRA after Q050, but diuretic prescriptions decreased from 82.7% to 76.1% (p<0.001). The remaining two subgroups who were assessed more than 3 months after diagnosis had no change in prescriptions. Overall, those diagnosed inpatient with a >3-month follow-up had the highest rates of HF medication use after Q050 billing code of all subgroups (73.9% RASi, 68.2% BB, 21.1% MRA, and 77.9% diuretics).

## Discussion

To our knowledge, this was the first study on the effect of a pay-for-performance program on HF process-of-care measures in primary care. Our novel findings were 1) there was a marginal increase in the overall prescriptions of heart failure medications after Q050 billing 2) diuretics were highly prescribed but disease-modifying therapies remained underutilized.

Since its inception, the Q050 billing code has been widely adopted by family physicians^10^. It provided clinical guidance and financial compensation to FPs for managing an increasingly complex disease. However, a previous analysis showed that the use of this billing code was not associated with any change in key clinical outcomes such as mortality, HF hospitalizations, and emergency department visits related to HF ^5^; our current analysis on process-of-care measure might explain the previously-observed outcomes. Prior studies showed that the use of disease-modifying therapies was associated with improved outcomes, yet we found that quarter of patients still were not prescribed these drugs even after the incentive billing code ^11^. In the largest and most sensitive subgroup of patients who were diagnosed inpatient and assessed by FPs within 3 months of diagnosis, the improvement in prescription rates was modest at best. Surprisingly, this subgroup also had the lowest rate of decongestive therapies, despite their presentation with acute HF. In comparison, the subgroup with the most comorbidities did receive the highest rates of HF prescriptions, yet they had a much-delayed follow-up with their family physicians despite being the “sickest” patients.

The literature on P4P in primary care is conflicting. In 2007, British Columbia established a P4P scheme for primary care physicians who manage patients with two or more chronic conditions. A subsequent evaluation showed that there was no increase in the amount of patient care or the continuity of care; in fact, the total rate of hospital admission increased relative to the pre-intervention trend ^12^. In contrast, P4P program for cancer screening in Ontario showed promising increase in mammogram and colorectal cancer screening ^13^. Extensive studies on the Quality and Outcome Framework, the world’s largest pay-for-performance scheme in primary care implemented in the United Kingdom in 2004, were similarly inconclusive ^14,15^.

Qualitative studies show that HF management in primary care is challenged by uncertainties around treatment options and benefits, underuse of clinical guidelines, limited access to specialist care, and difficulties with poly-pharmacies and comorbidity management ^16^. These challenges contribute to clinical inertia, a well-established phenomenon in the management of chronic conditions such as HF ^17,18^. While Q050 provided a modest financial incentive, it did not adequately address the various barriers to care. Currently, a randomized controlled trial is underway to assess whether more support for FPs – a medication titration plan, a HF helpline, a problem-solving guide – can improve processes of care, patient functional capacity, and patient-reported symptoms ^19^.

We are limited by the variables available in our databases. Not all prescriptions were filled and dispensed, and not all dispensed medications were taken by patients. While ODB adequately captured medication dispensation, it likely underestimated the true prescription rate and overestimated the true adherence rate. Secondly, we had no information on clinical data such as blood pressure or heart rate measurements. Information on left ventricular ejection fraction (LVEF) was not available and we could not assess HF therapies specific to the phenotypes of reduced and preserved ejection fraction. However, patients with HF often had comorbidities that warranted treatments with similar medications regardless of their HF phenotype; for example, we expected a higher rate of RASi prescription given that 88% of the cohort had hypertension. Lastly, we did not capture any changes in medication dosage, but clinical practice generally supports a shotgun approach to HF therapies rather than sequential initiation and dose escalation^8^. Our focus on the number of therapies was still a relevant marker of care.

In summary, pay-for-performance incentive in primary care management of HF had marginal effect on process-of-care measures. Further analyses of barriers to care are needed to understand clinical inertia and improve HF management in primary care.

## Data Availability

The datasets from this study are held securely in coded form at the ICES. Although legal data?sharing agreements prohibit ICES from making the data set publicly available, access may be granted to those who meet prespecified criteria for confidential access, available at https://www.ices.on.ca/DAS. The full dataset creation plan and underlying analytic code are available from the authors upon request, understanding that the computer programs may rely on coding templates or macros that are unique to ICES and are, therefore, inaccessible or may require modification.

## Acknowledgements

This study was supported by ICES, which is funded by an annual grant from the Ontario Ministry of Health (MOH) and the Ministry of Long-Term Care (MLTC). Parts of this material are based on data and information compiled and provided by CIHI and the Ontario Ministry of Health. The analyses, conclusions, opinions and statements expressed herein are solely those of the authors and do not reflect those of the funding or data sources; no endorsement is intended or should be inferred. We thank IQVIA Solutions Canada for use of their Drug Information File.

